# Peripheral Artery Disease and COVID-19 Outcomes: Insights from the Yale DOM-CovX Registry

**DOI:** 10.1101/2021.08.16.21262118

**Authors:** Kim G. Smolderen, Megan Lee, Tanima Arora, Michael Simonov, Carlos Mena-Hurtado

## Abstract

**Background:** Both COVID-19 infection and peripheral arterial disease (PAD) cause hypercoagulability in patients, and it remains unknown whether PAD predisposes patients to experience worse outcomes when infected with SARS-CoV-2.

**Methods:** The Yale DOM-CovX Registry consecutively enrolled inpatients for SARS-CoV-2 between March 1, 2020, and November 10, 2020. Adjusted logistic regression models examined associations between PAD and mortality, stroke, myocardial infarction (MI), and major adverse cardiovascular events (MACE, all endpoints combined).

**Results:** Of the 3,830 patients were admitted with SARS-CoV-2, 50.5% were female, mean age was 63.1 ±18.4 years, 50.7% were minority race, and 18.3% (n = 693) had PAD. PAD was independently associated with increased mortality (OR=1.45, 95% CI 1.11-1.88) and MACE (OR=1.48, 95% CI 1.16-1.87). PAD was not independently associated with stroke (p=0.06) and MI (p=0.22).

**Conclusion:** Patients with PAD have a >40% odds of mortality and MACE when admitted with a SARS-CoV-2, independent of known risk factors.

## Introduction

COVID-19 is a disease with many diverse manifestations, ranging from asymptomatic disease to mild symptoms, and more severely, thrombotic events, acute respiratory syndrome, and death. As our understanding for the disease evolved, it has become evident that widespread vascular coagulopathy and infection-triggered inflammatory escalations may be mechanisms that explain adverse outcomes such as respiratory failure and mortality.^1^

As community spread of infection continues to increase globally, understanding which subpopulations are at highest risk of adverse outcomes following SARS-CoV-2 infection is key. One common comorbidity that is highly prevalent and underdiagnosed in the overall population is peripheral artery disease (PAD). Hypercoagulation, part of the underlying pathogenesis of this atherosclerotic disease, is shared with the SARS-CoV-2 disease course.^2^ Given this underlying vulnerability and risk profile that these patients inherently carry, it is unknown whether infection with SARS-CoV-2 may additionally worsen the prognostic outcomes of patients with PAD who are infected with SARS-CoV-2.

We therefore aimed (1) to compare patient profiles in inpatients of COVID-19 by history of PAD status; and (2) compare outcomes (cardiac complications and mortality) by PAD status. We hypothesize that in patients admitted with COVID-19, patients with PAD disproportionally experience worse outcomes, including more thrombotic events and higher mortality compared to patients without PAD.

## Methods

The Yale DOM-CovX Registry is an observational longitudinal registry conducted at the Yale New Haven Health System. The registry consecutively enrolled all inpatients admitted to the Yale New Health System for SARS-CoV-2 between March 1, 2020, and November 10, 2020. Patients <18 years were excluded from the analyses. Data was queried from the electronic medical records for demographics, comorbidities, lab values, medications, procedures, vitals, and clinical outcomes. Key features such as death were validated through manual chart review for a random sampling of patients. This study was reviewed by the Yale Institutional Research Board and given the deidentified nature of the data, the research was deemed non-human subjects research.

History of PAD was derived from patients’ clinical history as documented in the medical records per the International Classification of Diseases 10^th^ revision (ICD-10). Patient demographics, medical history (Elixhauser comorbidity index),^3,4^ labs, and clinical outcomes were recorded. Comparisons by PAD status were tested through Chi-square tests for categorical variables, and student-t-tests for continuous variables or non-parametric tests, as appropriate.

To examine the association between PAD and COVID-19 outcomes, we used a logistic regression model for the endpoints mortality, major adverse cardiovascular events (MACE) (mortality, stroke, myocardial infarction (MI)), stroke individually, and MI individually, adjusting in stepwise models for (1) patient demographics (age, sex, race, ethnicity), (2) PAD risk factors (body mass index, smoking, lipid disorder, diabetes, hypertension), (3) history of coronary disease (chronic heart failure, myocardial infarction), and (4) maximum D-dimer.^5^

## Results

Out of a total of 3,830 patients that were admitted with SARS-CoV-2, 50.5% were female, with a mean age of 63.1 ±18.4 years, 50.7% were Non-white race, and 18.3% (n = 693) had PAD (Table 1). Patients with PAD vs. no PAD had a higher rate of mortality (24.5% vs. 12.7%, p<0.001), stroke (5.3% vs. 2.6%, p<0.001), MI (22.7% vs. 6.7%, p<0.001), and overall MACE (44.0% vs.19.2%, p<0.001) (Table 2).

**Table 1.** Patient Characteristics

|  | Total<br>(N = 3830) | PAD<br>(N = 693) | No PAD<br>(N = 3087) |
| --- | --- | --- | --- |
| Female Sex | 1909 (50.5) | 344 (49.6) | 1565 (50.7) |
| Age (mean ± SD) | 63.1 ± 18.4 | 72.5 ± 13.6 | 61.1 ± 18.6 |
| Race |  |  |  |
| Asian | 70 (1.9) | 6 (0.9) | 64 (2.1) |
| Black | 267 (26) | 214 (31.1) | 753 (24.8) |
| White | 1834 (49.3) | 382 (55.4) | 1452 (47.9) |
| Other/unknown | 837 (22.5) | 84 (12.2) | 753 (24.8) |
| Hispanic Ethnicity | 969 (25.6) | 107 (15.4) | 862 (27.9) |
| Comorbidity history |  |  |  |
| AFib/AFlutter | 605 (16) | 274 (39.5) | 331 (10.7) |
| Diabetes | 1040 (27.5) | 387 (55.8) | 653 (21.2) |
| CVD | 631 (16.7) | 263 (38.0) | 368 (11.9) |
| Chronic heart failure | 823 (21.8) | 392 (56.6) | 431 (14.0) |
| Chronic kidney disease | 827 (21.9) | 371 (53.5) | 456 (14.8) |
| COPD | 771 (20.4) | 293 (42.3) | 478 (15.5) |
| Hypertension | 1063 (28.1) | 482 (69.6) | 581 (18.8) |
| Lipid disorder | 2009 (53.1) | 615 (88.7) | 1394 (45.2) |
| Myocardial infarction | 441 (11.7) | 222 (32.0) | 219 (7.1) |

*Results are presented as numbers (percentages).*
*Abbreviations: AFib: atrial fibrillation, Aflutter: atrial flutter, COPD: chronic obstructive pulmonary disease, SD: standard deviation*

**Table 2.** Patient Outcomes

|  | Total<br>(N = 3830) | PAD<br>(N = 693) | No PAD<br>(N = 3087) | P-value |
| --- | --- | --- | --- | --- |
| Mortality | 562 (14.9) | 170 (24.5) | 392 (12.7) | <0.001 |
| Stroke | 117 (3.1) | 37 (5.3) | 80 (2.6) | <0.001 |
| Myocardial Infarction | 365 (9.7) | 157 (22.7) | 208 (6.7) | <0.001 |
| MACE | 898 (23.8) | 305 (44.0) | 593 (19.2) | <0.001 |

*Results are presented as numbers (percentages).*
*Abbreviations: MACE: major adverse cardiac events*

After adjusting for patient demographics, PAD risk factors, history of CAD, and maximum D-dimer, PAD was independently associated with an increased mortality risk (odds ratio (OR) = 1.45, 95% confidence interval (CI): 1.11-1.88) and MACE (OR=1.48, 95% CI: 1.16-1.87) (Figure 1). PAD was not independently associated with stroke (p=0.06) after adjusting for demographics (step 1) and MI (p=0.22) after adjusting for demographics, PAD risk factors, and history of CAD (step 3).

**Figure 1.**
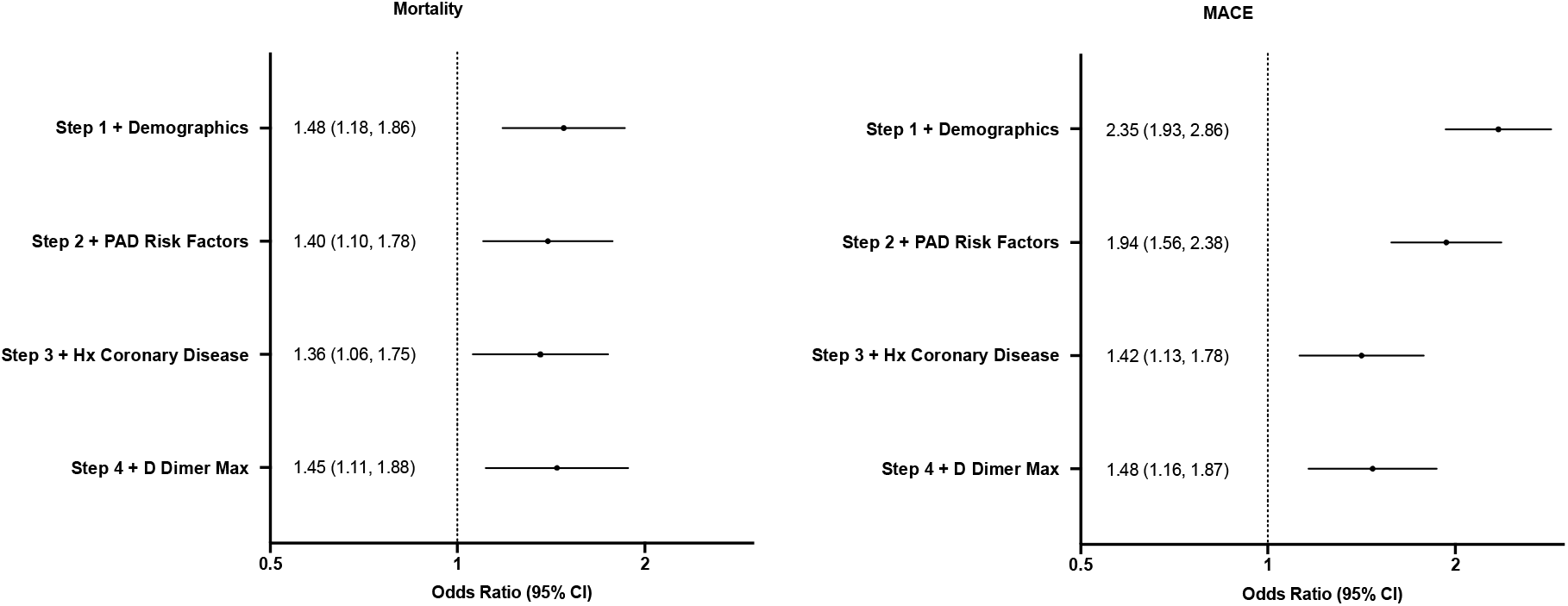
The association between peripheral artery disease and MACE in patients admitted for COVID-19 following sequential covariate adjustment Covariate adjustment: Step 1: age, sex, race, ethnicity; Step 2: body mass index, smoking, lipid disorder, diabetes, hypertension; Step 3: history of chronic heart failure, history of myocardial infarction; Step 4: D-Dimer Max. Abbreviations: MACE, Major Adverse Cardiovascular Events; PAD, peripheral artery disease; Hx, history; CI, confidence interval.

## Discussion

In inpatients with COVID-19, those with PAD have over a 40% increase in odds of both mortality and MACE, independent of known risk factors. Previous studies have shown a heightened risk of thromboembolic disease in patients with COVID-19,^1,5^ but to knowledge, our study is the first to look at outcomes specifically in patients with PAD. Despite most patients with COVID-19 receiving prophylactic heparin while inpatient, they continue to have high rates of thromboembolism.^1^ Future studies should look at specific pharmacologic strategies in patients with PAD to reduce the risk of MACE and mortality. Furthermore, understanding the risk profile for COVID-19 associated with PAD will also help us to inform patients with this vulnerability as to what their risks are and to maximize preventive strategies amongst those with the highest risk.

Our study’s limitations include a that our retrospective database review only included information encoded in patient medical records and had a possibility of underdiagnosed stroke, MI, or PAD. Furthermore, we encompassed a wide date range, with management strategies changing over time as the pandemic continued, for which our study design did not allow us to take these changes into account.

## Data Availability

Data is available upon request to the authors.

## Acknowledgements

The authors report no acknowledgements.

## Sources of Funding

This research received no specific grant from any funding agency in the public, commercial, or not-for-profit sectors.

## Disclosures

Dr. Smolderen reports grant support from Johnson & Johnson, Cardiva, Abbott, and she is a consultant for Optum Labs. Dr. Mena-Hurtado serves as a consultant for Abbott, Cook Medical, Medtronic, Cardinal Health, and Optum Labs LLC.

## Notes

### Funding Statement

No external funding for this work was received.

### Author Declarations

This study was reviewed by the Yale Institutional Research Board and given the deidentified nature of the data, the research was deemed non-human subjects research.

